# Association of the EEG Correlate Of Injury to the Nervous System (COIN) Index with Focal Cerebral Injury in Children Receiving Extracorporeal Membrane Oxygenation

**DOI:** 10.64898/2026.08.06.26359920

**Authors:** Rezvaneh Ghasemzadeh, Kathryn Finlay, Yi Li, Adam L. Numis, Rohit Jain, Edilberto Amorim, Giulia M. Benedetti, Craig A. Press, Dana B. Harrar, Ajay X. Thomas, Loren D. Sacks, Christine K. Fox, Mauro Caffarelli

**Author notes:** Corresponding Author: Mauro Caffarelli, MD, 675 Nelson Rising, NS 402, San Francisco, CA 94158.

## Abstract

**BACKGROUND:** Children receiving extracorporeal membrane oxygenation (ECMO) are at high risk for focal cerebral injury (FCI). There is emerging evidence that electroencephalography (EEG) may aid FCI detection. The EEG Correlate of Injury to the Nervous System (COIN) index quantifies and displays focal background asymmetries. We evaluated whether COIN is associated with FCI in pediatric ECMO.

**METHODS:** Retrospective, cross-sectional study of patients age 28 days to 21 years, on venoarterial ECMO at a tertiary children’s hospital, who received EEG monitoring and neuroimaging during ECMO. COIN was calculated from all available EEG data. COIN of 0 implies a symmetric EEG and negative COIN values are observed with FCI. Median COIN values near FCI recognition time were compared to median COIN values from randomly selected control EEG batches using logistic regression. A receiver operator characteristic curve was used to identify multilevel FCI test ranges. Likelihood ratios were calculated to estimate the posttest FCI probability for each COIN range.

**RESULTS:** During the 8-year study period (2015-2023), 33 of 142 ECMO runs met study criteria for COIN analysis. Twelve patients (36%) had FCI. The COIN cutoff of –13.3 had 92% sensitivity and 67% specificity for FCI. The COIN cutoff of –27.7 had 67% sensitivity and 90% specificity. Likelihood ratios were 0.13 for COIN (0 to –13.3), 1.1 for COIN (–13.3 to –27.7), and 7.0 for COIN (< –27.7). Posttest probability was 0.02, 0.13, 0.49 in each respective range.

**CONCLUSION:** FCI on ECMO is associated with COIN-measured EEG asymmetry. COIN may support FCI risk-stratification during ECMO.

## INTRODUCTION

Children on extracorporeal membrane oxygenation (ECMO) are at high risk for focal cerebral injury (FCI).^1–3^ FCI – which includes arterial ischemic stroke, parenchymal hemorrhage, and subdural hemorrhage – occurs in approximately 12% of ECMO runs with a 21% absolute mortality increase and major neurodevelopmental morbidity in 30% of survivors at 1-year follow up.^4^ ECMO utilization has quadrupled in children in the last two decades, yet rates of FCI morbidity and mortality are largely unchanged.^5–8^ Timely FCI diagnosis may enable mitigating interventions, but clinical symptom recognition is often impeded by sedation and neuromuscular blockade, and obtaining imaging on ECMO can be difficult and impose patient risk. Such examination limitations may be overcome with electroencephalography (EEG) to detect background asymmetries suggestive of FCI and aid in patient selection for imaging.^3,9^

While EEG monitoring can inform the risk of FCI,^6^ it is currently driven by intermittent qualitative interpretation of raw tracings by experts.^10^ Quantitative EEG, which performs mathematical transformations to allow for numeric and colorimetric display of EEG data, may support detection of FCI during pediatric ECMO,^9,11^ especially in high risk periods such as circuit changes.^12^ Our group developed the quantitative EEG-based Correlate of Injury to the Nervous System (COIN) index to support non-expert recognition of asymmetric EEG background abnormalities suggestive of FCI.^13^

COIN measures and displays focal EEG power attenuation in the theta (4-8 Hz) and alpha (8-12 Hz) power bands. COIN quantifies focal power attenuation on a bipolar EEG montage as a single continuous variable where ‘0’ implies a symmetric EEG and increasingly negative values are observed with focal power attenuation in one or more bipolar channels. Prior exploratory work has shown that COIN values below –20 are associated with large arterial ischemic stroke and intracranial hemorrhage with the potential to aid in bedside FCI detection.^13–15^ Here, we used retrospective clinically-obtained EEG data during ECMO at a single institution to explore our hypothesis that negative COIN values are associated with FCI that occurs during ECMO.

## METHODS

### Study design and population

This is a retrospective, cross-sectional cohort study of pediatric patients supported with venoarterial ECMO at a quaternary children’s hospital from January 1^st^, 2015, to December 31^st^, 2023. The inclusion criteria were: 1) Patients aged 28 days to 21 years; 2) Neuroimaging performed during ECMO or up to two months post-decannulation; and 3) EEG recording during ECMO. Cerebral magnetic resonance imaging, head computed tomography or, for children aged <12 months old, head ultrasound, were all deemed acceptable neuroimaging modalities. Patients monitored using a limited neonatal EEG montage (typically <1 month corrected post-gestational age) were excluded. A waiver of consent for minimal risk for the study (IRB #20-30126) was approved on April 13, 2020, by the University of California’s Institutional Review Board.

### Clinical Data Collection

The University of California San Francisco institutional ECMO registry data was used to identify patients for inclusion and collect data variables (routinely collected for the Extracorporeal Life Support Organization (ELSO) compliance) including patient sex, age at time of ECMO cannulation, ECMO indication (extracorporeal cardiopulmonary resuscitation [ECPR], cardiac, or pulmonary), site of arterial cannulation, duration of ECMO run, and survival to discharge. ELSO Registry data were collected and prepared for analysis by our hospital ECMO nurse coordinator. Chart review was performed in included patients to generate a supplemental data registry including additional variables: ECMO circuit change times, EEG start and stop time, qualitative EEG results (i.e., focal seizures or focal background abnormalities consisting of unilateral focal slowing, focal voltage attenuation, or voltage asymmetry), imaging data, and FCI findings with onset time data. Because the cohort size was small, case report forms were screened for missing or mis-entered data and validated by the study authors (RG and MC).

### Primary Outcome: Focal Cerebral Injury (FCI)

FCI was radiographically defined as lateralized ischemia or hemorrhagic parenchymal brain injury greater than 1cm^3^, or as subdural hemorrhage causing midline shift.^12^ Patient charts were initially screened for clinical neuroimaging reports suggestive of FCI. Reports describing study criteria for FCI were validated by a pediatric neuroradiologist (YL).

The time of clinical FCI recognition was obtained via chart review as described in a prior report.^12^ The documented clinical signs used for defining FCI were one or more of the following: 1) new clinical examination abnormality (e.g., pupillary changes, lateralized weakness, rhythmic extremity movements); 2) when available, new EEG abnormality (i.e., unilateral focal slowing, focal voltage attenuation, focal seizures, or voltage asymmetry); or 3) any other new clinical findings leading clinicians to investigate with head imaging. In instances when FCI was an incidental finding on imaging, the time of the neuroimaging report was used as the time of documentation of FCI.

### EEG Data Collection and COIN calculation

All EEG data recorded during ECMO and up to 48 hours after decannulation were identified and exported from Natus (Natus Medical Inc., Middleton, WI) as European data format (.edf) files. COIN was calculated from raw EEG datafiles on MATLAB (MathWorks, Natick, MA) per our previously reported algorithm.^13^ Files were deidentified and imported into using the open source FieldTrip package.^16^ FieldTrip algorithms were used to preprocess and filter data to include frequencies between 1-70 Hz with a notch filter of 60 Hz.

A bipolar montage was applied, and the study was divided into consecutive nonoverlapping 4-second trials. Artifact detection and rejection was performed using algorithms in Fieldtrip to detect clipping artifact (amplitude threshold 0.05 uV) and threshold artifact (>300 uV) with z-score rejection for muscle artifact (50-99 Hz, z-threshold=4) and movement artifact (1-5 Hz, z-threshold=4). Trials containing <2.5s of artifact had the periods of artifact removed from the trial with remaining EEG data from the trial used for analysis. Trials containing artifact in >2.5s of the trial were rejected.^15^

Cleaned EEG data were passed through a fast-Fourier transform to yield frequency power data for every 4-second trial in each bipolar channel. COIN values are calculated for each EEG channel, and channels with negative values are summed to generate a summary COIN value per 4-second trial. Consecutive datapoints are labelled with a timestamp which is used to generate trendlines relative to the time of clinical FCI recognition or ECMO start. COIN values from trials over an 8-hour epoch are used to calculate a median COIN value.

Epoch selection was handled differently for FCI and controls. To avoid sampling of EEG prior to FCI onset, we sampled epochs spanning the first available 8 hours of EEG data, starting as early as 3 hours before clinical FCI symptom recognition through as late as 24 hours after clinical FCI symptom recognition. COIN visualizations were generated for comparison to available neuroimaging in patients with FCI.

Because this was an exploratory analysis of large-volume data, EEG studies were not read to obtain timestamped seizure start and stop times or sedative bolus doses. To reduce the effect of non-timestamped clinical events that may affect the background EEG in control patients, we sampled 10 random 8-hour epochs to obtain 10 separate median COIN values per patient. The random epoch sampling was not configured to prevent overlapping EEG data sampling, which was necessary for patients with shorter recording periods. This resulted in 10 separate sets of control COIN samples which were used for statistical analysis.

### Statistical Analysis

To assess for selection bias, clinical features of children on venoarterial ECMO meeting age criteria were compared based on whether they met remaining criteria for COIN analysis using t-test for proportions, Mann-Whitney U test for medians, and t-test for means. Within the COIN-analyzed group, Median COIN values were compared to FCI status using logistic regression. Sensitivity and specificity were calculated across a range of COIN thresholds using the FCI COIN values against the 10 batches of control COIN values. Final reported values were calculated using the mean of 10 median control values for each patient.

We sought to assess COIN’s performance as a multi-level test. Estimated sensitivity and specificity values were used to generate a receiver operator characteristic (ROC) curve. The ROC was used to identify two COIN cutoffs for multilevel test ranges that maximized positive and negative Likelihood Ratios (LRs) while maintaining balance in sample distribution across the three multilevel ranges. COIN-range based LRs were used to calculate post-test probability of FCI using a literature-reported FCI incidence of 12%.^4^ Selected covariates (age, sex, ECMO indication, mortality, cannulation sites, ECLS duration, focal EEG background attenuation, focal seizures, FCI type) were compared between patients in each multi-level test range to assess for potential sources of bias.

## RESULTS

During the study period, there were 142 ECMO runs in 132 patients, of which 88 were venoarterial and met age criteria. Of these, 37 runs in 37 unique patients received head imaging and full montage EEG. FCI was identified in 16 patients, 4 of whom were excluded from analysis because they did not have EEG recorded near the time of FCI symptom recognition (3 hours before to 24 hours after). The final COIN-analyzed cohort consisted of 12 (36%) patients with FCI and 21 (64%) patients without FCI. The 12 patients with FCI were diagnosed with arterial ischemic stroke (AIS) in 9 (75%), subdural hemorrhage with midline shift in 2 (17%), parenchymal hemorrhage in 1 (8%). The 21 remaining patients were analyzed as controls, resulting in a total population of 33 patients for COIN analysis. ECMO indication in children meeting criteria for COIN analysis was more likely to be ECPR and less likely to be cardiac. Consort flow diagram and tabulated comparison of clinical characteristics between patients are shown in the supplemental materials.

The COIN-analyzed study population had a median age of 0.44 (0.30 – 3.9) years and consisted of 12 (36%) females. The ECMO indication was cardiac in 10 (30%), pulmonary in 2 (6%), and ECPR in 21 (64%). Arterial cannulation site was aorta in 9 (27%), carotid artery in 23 (70%), and femoral artery in 1 (3%). Hypoxic ischemic brain injury was diagnosed in 8 (24%) children: 3 with FCI and 5 without FCI. Six (50%) children with FCI were deceased at hospital discharge, and 8 (38%) children without FCI were deceased at discharge. There was not a statistically significant difference in death rates between children with and without FCI (p = 0.76).

### COIN Analysis

COIN was calculated from all available EEG data in the 33 study patients. Figure 1 shows calculated COIN values and visualizations from a patient with AIS diagnosed six hours after an ECMO circuit change. Sample 4-second EEG trials from before, during, and after the circuit change demonstrate the clear development of electrographic asymmetry. The asymmetry is reflected in a progressive decline in the COIN trendline with visualization of power attenuation (blue shading) in multiple left-hemispheric channels.

**Figure 1:**
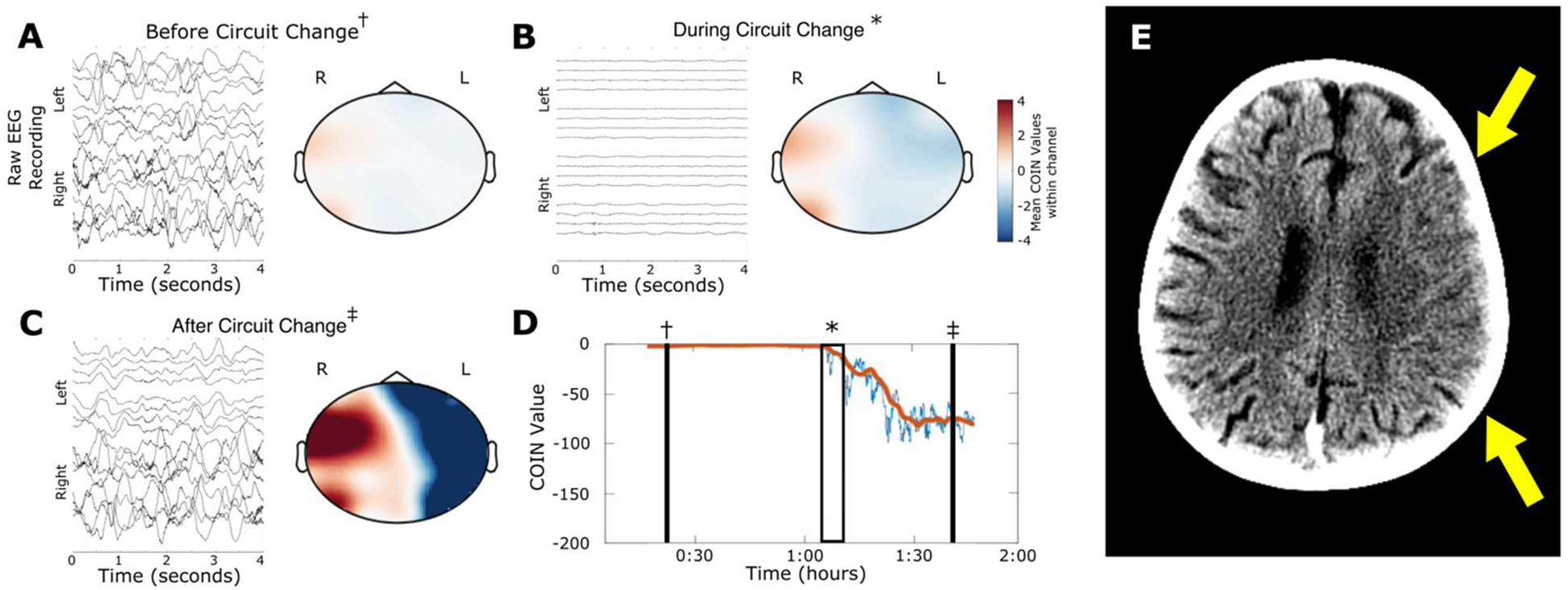
Example of COIN output with FCI sustained during ECMO circuit change. Sample 4-second EEG trials and corresponding COIN visualizations are shown before (A), during (B), and after (C) circuit change. Summary COIN trendline (D) shows a stable trendline at 0 two hours prior to the circuit change, with a downward progression to –80 over the two hours following circuit change. Computed tomography 6 hours after circuit change shows effacement of sulci and gray-white border in left middle cerebral artery territory (E).

Figure 2 shows swimmerplots of continuous COIN trendlines in all FCI and non-FCI patients. FCI swimmerplots are aligned based on the time of clinical FCI symptom recognition, whereas control swimmerplots are aligned based on ECMO start time. Swimmerplots in FCI show qualitatively depressed COIN values surrounding the time of FCI recognition, whereas swimmerplots in non FCI patients shows a mixture of patients with high COIN values (close to 0) and intermittent periods of depressed COIN values. Topographic COIN visualization aligned with radiographic FCI localization in 11 (92%) of the 12 patients with FCI. Figure 3 shows neuroimages alongside COIN visualization in patients with FCI.

**Figure 2:**
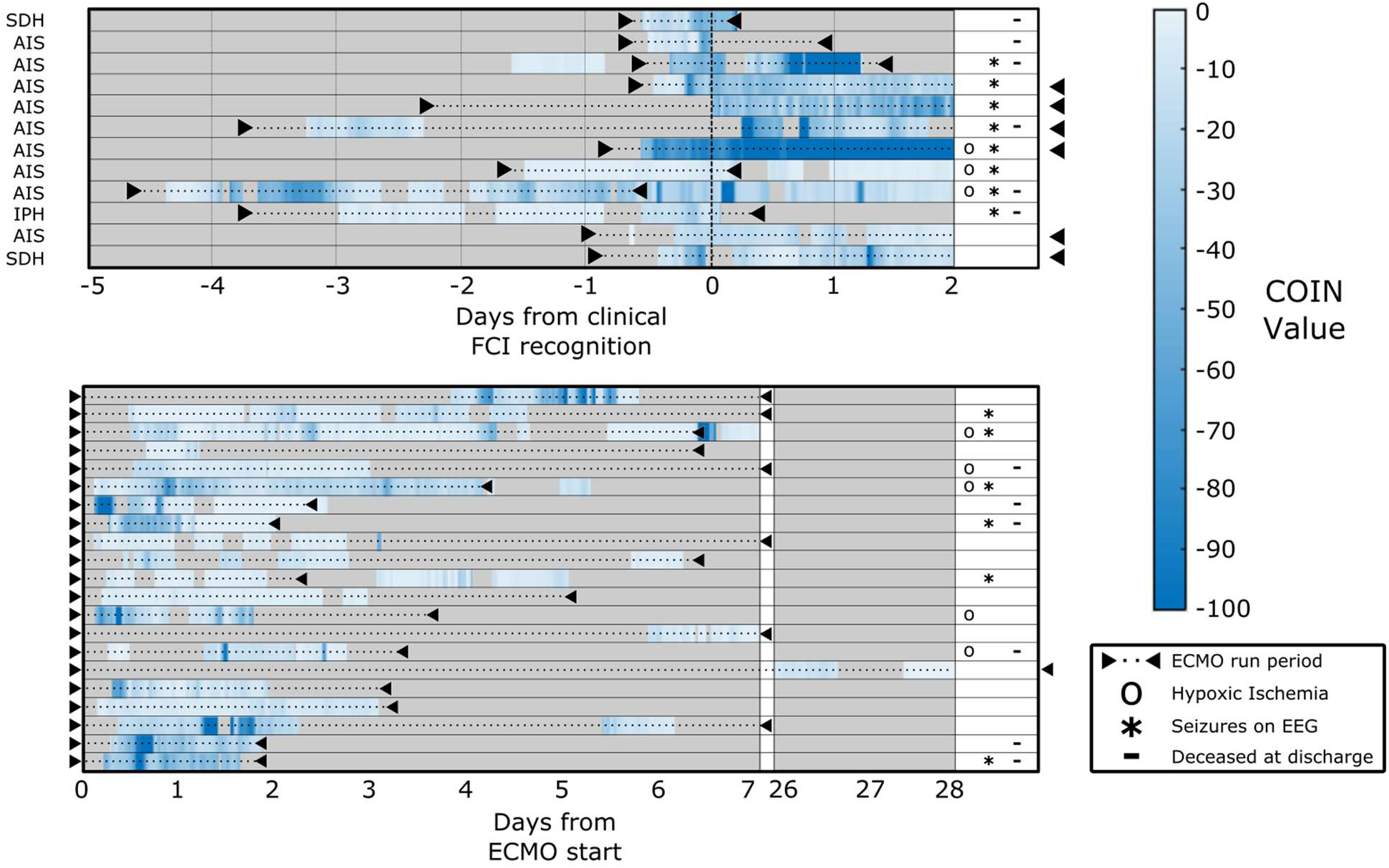
Swimmerplots of COIN trends over multiple days of EEG recording during ECMO in all patients with FCI (A) and without FCI (B). Time axis in FCI group is synchronized to the time of FCI symptom recognition as timepoint ‘0’ to enable qualitative view of COIN values before and after FCI recognition. Non-FCI time axis is synchronized to the ECMO start time. COIN values below –100 are saturated to the color associated with –100. Triangles and black dotted line indicate the duration of the ECMO run. Right-hand column shows patients with: Neuroimaging-reported hypoxic ischemic injury ‘o’; Electrographic seizures ‘*’ and; Deceased at discharge ‘-‘. AIS, arterial ischemic stroke; COIN, correlate of injury to the nervous system; ECMO, extracorporeal membrane oxygenation; EEG, electroencephalogram; FCI, focal cerebral injury; IPH, intraparenchymal hemorrhage; SDH, Subdural hemorrhage.

**Figure 3:**
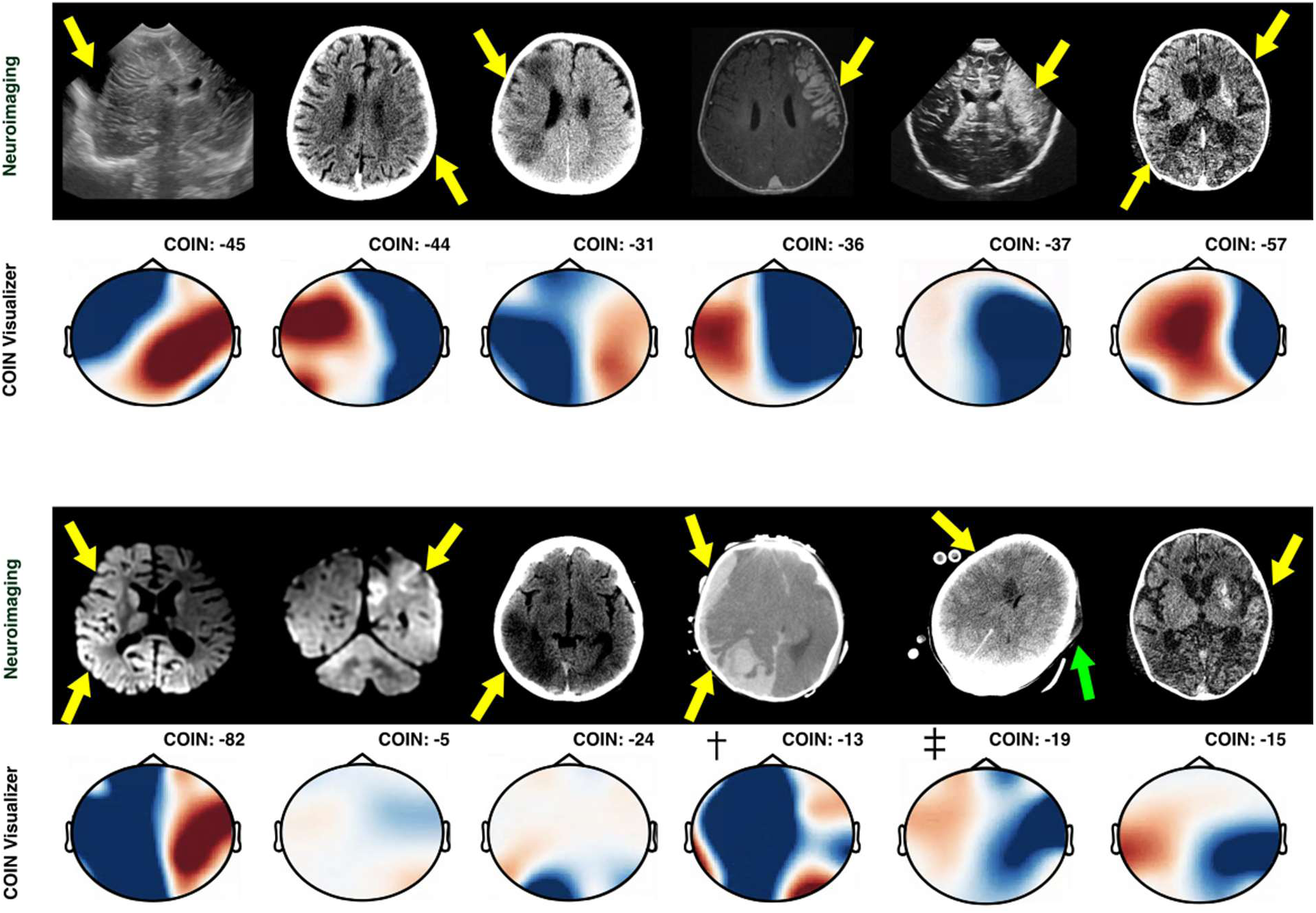
Visual comparison of neuroimaging and COIN topographic display with median COIN values in the 12 patients with FCI. Radiographic images are shown with patient’s right side on the left per standard convention, with COIN display mirroring radiographic lateralization. Cases from top-left to bottom-right follow the same order as in Figure 2. Yellow arrows indicate the localization of FCI. † Background EEG showed rapid onset right hemispheric attenuation with clinical herniation syndrome over one hour (hypertension, bradycardia, dilated nonreactive pupil); EEG was discontinued and care was withdrawn after neuroimaging diagnosis. ‡ Right caudate infarct (yellow arrow) was identified after qualitative clinical recognition of left hemispheric voltage attenuation; imaging additionally notable for left head rotation with left dependent scalp edema (green arrow) which may explain COIN findings. Image interpretations by the study neuroradiologist (YL) are available in the supplementary materials. COIN, correlate of injury to the nervous system; EEG, electroencephalography; FCI, focal cerebral injury.

The results of logistic regression and ROC analysis across median COIN values are shown in Figure 4. COIN values of –13.3 and –27.7 were identified at multi-level cutoffs (low risk, 0 to –13.3, n=16; medium risk, –13 to –27.7, n=8; high risk, –27.7 and below, n=9). The COIN cutoff of –13.3 was associated with an overall OR of 22.0 (95% CI: 3.3 – 445.8), sensitivity of 92% (62% – 100%), and specificity of 67% (43% – 85%). The COIN cutoff of –27.7 was associated with an overall odds ratio of 19.0 (95% CI: 3.4 – 165.0), sensitivity of 67% (35% – 90%), and specificity of 90% (70% – 99%).

**Figure 4:**
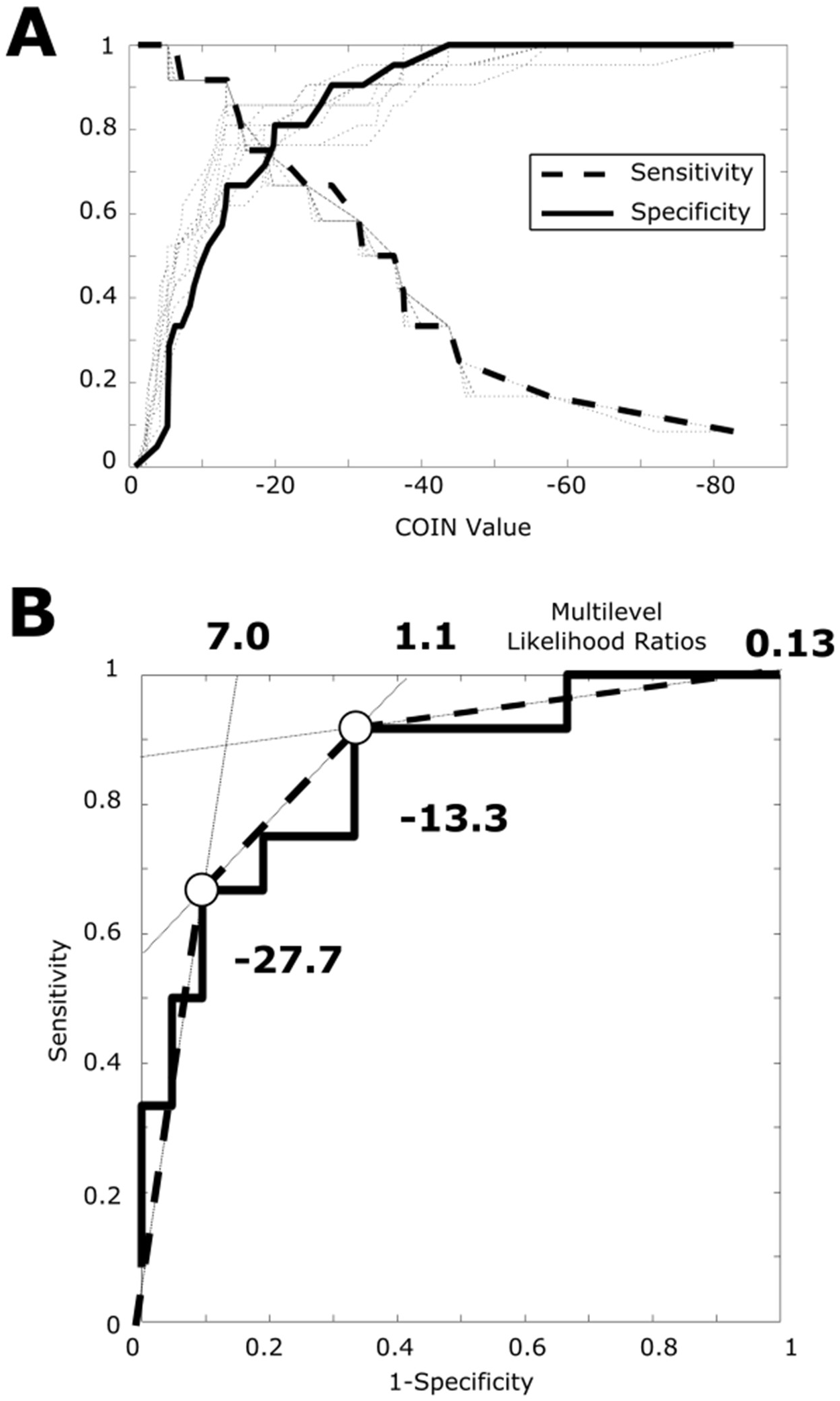
Results of logistic regression and ROC analysis. Measured sensitivity and specificity across all measured COIN values in the study cohort (A) across 10 control sample batches (light dotted lines) and pooled means (dark solid and dashed lines). Receiver operator characteristic curve (B) demonstrating sensitivity and specificity values for optimized multilevel cutoffs of – 13.3 and –27.7. The slope of the lines reflect the likelihood ratios of the resulting multilevel ranges. COIN, corelate of injury to the nervous system; ROC, receiver operator characteristic.

Estimated LRs in the different ranges were: 0.13 in the low-risk range, yielding a post-test FCI probability of 0.02 (with a pre-test probability of 0.12); 1.1 in the medium-risk range, yielding a post-test FCI probability of 0.13; and 7.0 in the high-risk range, yielding a post-test FCI probability of 0.49. Clinical characteristics, FCI status, and other post-test metrics are grouped by multi-level test range and summarized in Table 1.

**Table 1:**
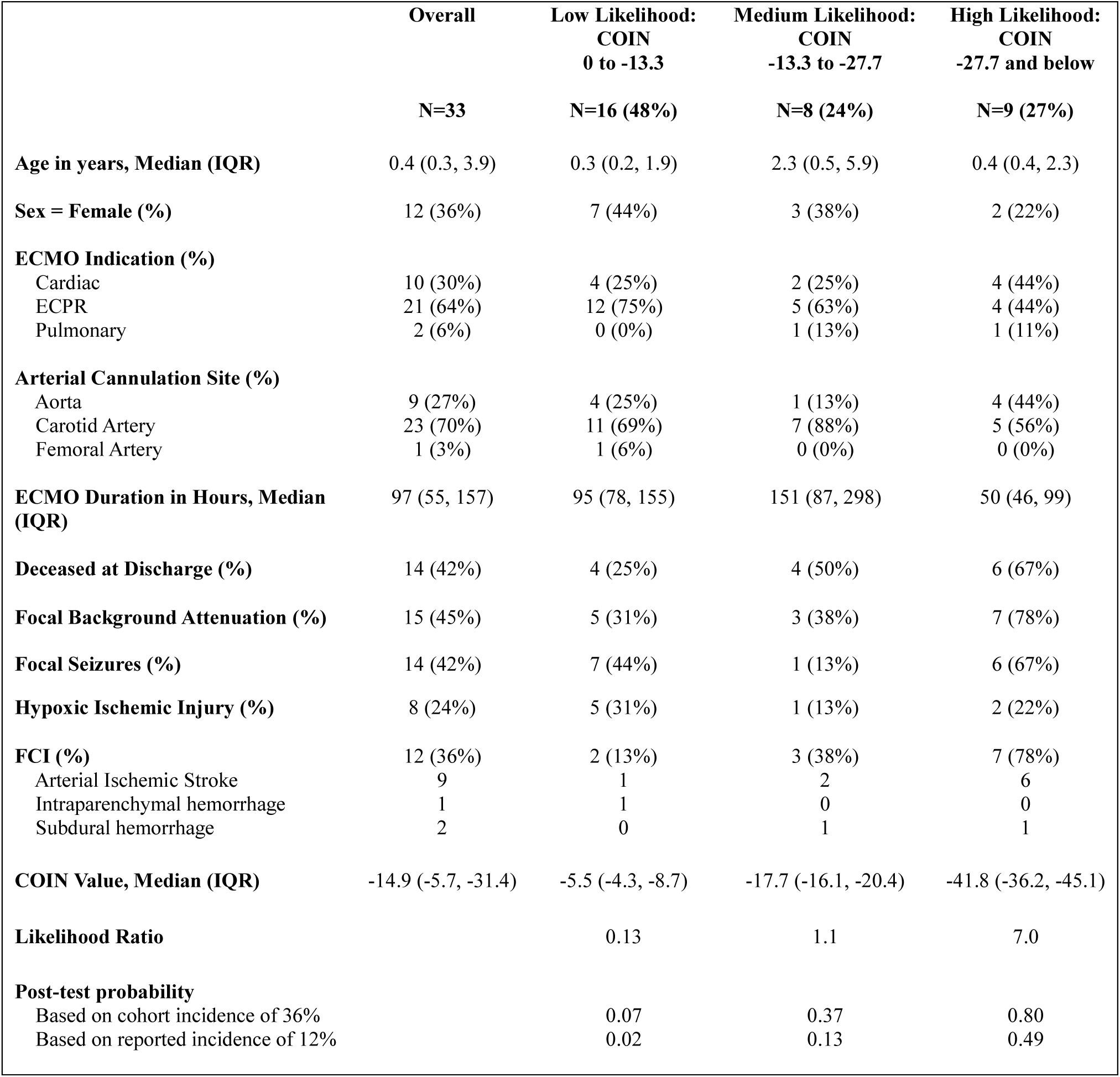
Baseline characteristics grouped by multi-level COIN ranges. Percentages in the Overall column are calculated using n=33, percentages in the multilevel range columns are calculated across rows using the overall count as the denominator. COIN: Correlate Of Injury to the Nervous system, ECMO: Extracorporeal Membrane Oxygenation, ECPR: Extracorporeal Cardiopulmonary Resuscitation, FCI: Focal Cerebral Injury, IQR: Interquartile Range, SD: Standard Deviation.

|  | **Overall** | **Low Likelihood:**<br>**COIN**<br>**0 to -13.3** | **Medium Likelihood:**<br>**COIN**<br>**-13.3 to -27.7** | **High Likelihood:**<br>**COIN**<br>**-27.7 and below** |
| --- | --- | --- | --- | --- |
|  | **N=33** | **N=16 (48%)** | **N=8 (24%)** | **N=9 (27%)** |
| **Age in years, Median (IQR)** | 0.4 (0.3, 3.9) | 0.3 (0.2, 1.9) | 2.3 (0.5, 5.9) | 0.4 (0.4, 2.3) |
| **Sex = Female (%)** | 12 (36%) | 7 (44%) | 3 (38%) | 2 (22%) |
| **ECMO Indication (%)** |  |  |  |  |
| Cardiac | 10 (30%) | 4 (25%) | 2 (25%) | 4 (44%) |
| ECPR | 21 (64%) | 12 (75%) | 5 (63%) | 4 (44%) |
| Pulmonary | 2 (6%) | 0 (0%) | 1 (13%) | 1 (11%) |
| **Arterial Cannulation Site (%)** |  |  |  |  |
| Aorta | 9 (27%) | 4 (25%) | 1 (13%) | 4 (44%) |
| Carotid Artery | 23 (70%) | 11 (69%) | 7 (88%) | 5 (56%) |
| Femoral Artery | 1 (3%) | 1 (6%) | 0 (0%) | 0 (0%) |
| **ECMO Duration in Hours, Median (IQR)** | 97 (55, 157) | 95 (78, 155) | 151 (87, 298) | 50 (46, 99) |
| **Deceased at Discharge (%)** | 14 (42%) | 4 (25%) | 4 (50%) | 6 (67%) |
| **Focal Background Attenuation (%)** | 15 (45%) | 5 (31%) | 3 (38%) | 7 (78%) |
| **Focal Seizures (%)** | 14 (42%) | 7 (44%) | 1 (13%) | 6 (67%) |
| **Hypoxic Ischemic Injury (%)** | 8 (24%) | 5 (31%) | 1 (13%) | 2 (22%) |
| **FCI (%)** | 12 (36%) | 2 (13%) | 3 (38%) | 7 (78%) |
| Arterial Ischemic Stroke | 9 | 1 | 2 | 6 |
| Intraparenchymal hemorrhage | 1 | 1 | 0 | 0 |
| Subdural hemorrhage | 2 | 0 | 1 | 1 |
| **COIN Value, Median (IQR)** | -14.9 (-5.7, -31.4) | -5.5 (-4.3, -8.7) | -17.7 (-16.1, -20.4) | -41.8 (-36.2, -45.1) |
| **Likelihood Ratio** |  | 0.13 | 1.1 | 7.0 |
| **Post-test probability** |  |  |  |  |
| Based on cohort incidence of 36% |  | 0.07 | 0.37 | 0.80 |
| Based on reported incidence of 12% |  | 0.02 | 0.13 | 0.49 |

## DISCUSSION

In this exploratory study, we demonstrate that focal EEG power attenuation quantified by COIN is associated with FCI in children and young adults on ECMO. COIN’s nature as a single continuous variable enables its characterization as a multi-level test with powerful risk stratification capability. Our results suggest that low COIN values between 0 and –13.3 nearly rules out FCI, whereas greater COIN values beyond –27.7 may carry a higher FCI risk and warrant further neurologic evaluation. Depending on the clinical context and specific clinical needs, COIN might provide an objective FCI risk screening tool in patients on ECMO with a limited neurologic exam.

FCI in ECMO results in high morbidity and mortality;^4,17^ this may be exacerbated by diagnostic delay. Typically, recognition of new FCI often does not occur until there is a clinically evident herniation syndrome or the patient develops seizures.^12^ In such cases, the presumption is that the FCI occurred many hours prior to the clinical manifestation. The delayed manifestation of FCI symptoms prevents benefit from mitigating interventions, such as pausing anticoagulation for hemorrhage or augmenting blood pressure for ischemia.^18,19^ Mechanical thrombectomy for thromboembolic large vessel occlusion stroke in ECMO has been described in adults and may be feasible in children with early recognition of occlusion onset.^20,21^

EEG markers of FCI — such as focal slowing and asymmetric voltage attenuation — are well described in large ECMO cohorts and are considered important adjuncts for FCI detection.^9,11^ While EEG during ECMO is primarily used to inform seizure detection and treatment,^8^ recent neonatal neuromonitoring guidelines in ECMO suggest EEG background assessment for FCI recognition.^7^ Current guidelines for EEG-based ischemia recognition rely on trained expert interpretation, which can only occur intermittently.^22^ While quantitative EEG metrics such as the alpha-delta ratio or brain symmetry index may support FCI detection,^9,11^ they are not amenable to threshold-based risk stratification for bedside screening.^13^ While this initial appraisal of COIN’s association FCI shows promise, more work is needed to understand how the dynamic behavior of COIN and other quantitative EEG metrics over time can be used to recognize FCI onset or progression.

This study’s design contains numerous limitations and biases that should be addressed via a larger validation cohort or prospective study design. The most notable limitation to this study is the reliance on clinically indicated EEG and head imaging for patient selection, which resulted in a large drop out of cases. While this mostly resulted in non-inclusion of patients without clinically-evident FCI, patients with clinical herniation symptoms may not receive EEG prior to discontinuation of care, and thus were included in this study design.^23^ Another limitation was the use of FCI-insensitive head ultrasound, which was necessary for expansion of our study population but also led to overrepresentation of infants as it can only be done in patients with an open fontanel. False-negative head ultrasound may have miscategorized FCI as non-FCI.^24^ Finally, the use of neuroimaging as the gold-standard reference for neurophysiologic ischemia detection is limited by the time-delay between FCI symptom recognition and image timing.^12^ We measured COIN values near the time of clinical FCI recognition, rather than imaging. Imaging that was delayed from FCI recognition time may have reflected interval worsening in FCI. Conversely, imaging in controls may have occurred before an FCI that occurred during EEG but was not clinically recognized. All these hypothetical possibilities will have biased our results toward the null.

Despite the study’s limitations, our findings have important implications for future research. Our small cohort size risks overfitting of test range cutoffs; expansion to include multi-center cohort data will enable validation while addressing many of the above biases, as different institutions have different EEG monitoring and head imaging practices.^25^ A larger FCI group will also enable study of how dynamic changes in COIN over time may support FCI recognition. Prospective evaluation of COIN with protocolized EEG monitoring during high risk epochs, such as circuit changes or decannulation,^12^ may elucidate how COIN can be clinically utilized as a marker of FCI risk. The ability to rapidly recognize FCI onset and streamline downstream clinical management would support better outcomes in this pediatric ECMO, where brain injury remains a serious and challenging problem.

## CONCLUSION

COIN values between 0 and –13.3 is associated with a low likelihood of FCI, whereas COIN below –27.7 is associated with a high likelihood of FCI. Further research is needed to validate COIN and determine how it may support early FCI recognition during ECMO.

## SOURCES OF FUNDING

Dr. Caffarelli is supported by the UCSF Clinical and Translational Science Institute, UCSF Catalyst Program, Hellman Society, Pediatric Epilepsy Research Foundation, and the Child Neurology Career Development Program (CNCDP) via the Kennedy Krieger Institute.

## DISCLOSURES

Dr. Caffarelli and Dr. Jain are founders of NeuroSentry, Inc. a producer of EEG software for brain injury detection.

## Data Availability

De-identified clinical and processed EEG metric data will be made available upon reasonable request,

## ABBREVIATIONS

COIN: Correlate Of Injury to the Nervous system index
ECMO: Extracorporeal Membrane Oxygenation
ECPR: Extracorporeal Cardiopulmonary Resuscitation
EEG: Electroencephalography
FCI: Focal Cerebral Injury
LR: Likelihood Ratio
ROC: Receiver Operator Characteristic

## Notes

### Competing Interest Statement

The authors have declared no competing interest.

### Author Declarations

A waiver of consent for minimal risk for the study (IRB #20-30126) was approved on April 13, 2020, by the University of California's Institutional Review Board

